# Risk Factors for Spontaneous Preterm Birth are Mediated through Changes in Cervical Length

**DOI:** 10.1101/2023.04.20.23288082

**Authors:** Hope M. Wolf, Shawn J. Latendresse, Jerome F. Strauss, Adi L. Tarca, Roberto Romero, Sonia S. Hassan, Bradley T. Webb, Timothy P. York

## Abstract

Although short cervical length in the mid-trimester of pregnancy is a one of the strongest predictors of preterm birth (*i.e*., parturition before 37 completed weeks), there is limited understanding of how the dynamics of cervical remodeling (*i.e*., changes in cervical length) leading up to labor and delivery can inform obstetrical risk. In this study, latent growth curve analysis was applied to serial cervical length measurements across pregnancy (median of 6; IQR = 3-8) to quantify characteristics of cervical change in a cohort of 5,111 singleton pregnancies consisting predominantly of Black women. A conditional mediation model including nine common maternal risk factors for spontaneous preterm birth as exogenous predictors accounted for 26.5% of the variability in gestational age at delivery (*P* < 0.001). This model provides insight into distinct mechanisms by which specific maternal risk factors influence preterm birth. For instance, effects of maternal parity and smoking status were fully mediated through cervical change parameters, whereas the influence of previous preterm birth was only partially explained, suggesting alternative pathways could be involved. This study provides the first account of the intermediary role of cervical dynamics in associations between known maternal risk factors and gestational age at delivery.

## INTRODUCTION

Preterm birth is the leading cause of neonatal morbidity and mortality worldwide (1), and babies born prematurely are at higher risk for long-term health issues, such as intellectual, developmental, and physical disabilities (1,2). A single mid-trimester cervical length measurement by transvaginal ultrasound is the best available technique for predicting (3–18) and preventing preterm birth, when paired with the administration of vaginal progesterone or cerclage in patients with a short cervix (18–41). Despite promise as a diagnostic tool this measure is characterized by relatively low sensitivity (9,42). Improvements in the predictive value may be achieved by taking into account the dynamic nature of cervical change across gestation (42–46) and consideration of maternal characteristics (47–50).

The length of the cervix, defined as the distance between the internal and external ors of the cervical canal, is easily measured by transvaginal ultrasonography over the course of pregnancy (47,51). Estimates for the mean length of the cervix in the mid-trimester vary between 35 and 45 mm, depending on the population (52–54), with cervical lengths shorter than 25 mm before 24 weeks meeting the clinical definition of a short cervix (28). A short cervix in the mid-trimester is associated with a six-fold increase in the risk of preterm delivery (4), and the earlier in pregnancy that the shortening occurs, the higher the risk for spontaneous preterm birth (3–18).

Cervical shortening is often thought of as the initial step of cervical ripening leading to the onset of labor and can begin several weeks before parturition (44–46,55–65). In most uncomplicated full-term pregnancies, the length of the cervix remains constant or gradually decreases beginning around 30 weeks (66,67). However, in some pregnancies rapid and progressive cervical shortening can occur earlier (46,66,68,69), which is associated with an increased risk for many adverse pregnancy outcomes, including preterm premature rupture of membranes (pPROM) (70,71), ascending infection (72–74), preterm labor (75–77), and premature delivery (3–16,18).

Maternal characteristics, such as advanced maternal age (53), BMI > 25 (78,79), higher parity (48), Black race/ethnicity (80), tobacco use (81), and history of cervical trauma (82) have all been associated with an increased risk of a short cervix in the mid-trimester. In a recent study by Gudicha et al. an adjustment of the threshold to define a short cervix in the mid-trimester or later in gestation to account for maternal characteristics resulted in a substantial improvement in the sensitivity for prediction of women at risk for spontaneous preterm delivery (35). Thus, insight into the mechanism describing how maternal risk factors contribute to cervical shortening across pregnancy may provide an understanding on how maternal risk factors influence preterm birth (69,83,84).

This study aimed to elucidate the etiological relationships between common maternal risk factors for preterm birth and the physiological changes in cervical length occurring during pregnancy. A multi-stage analytic strategy was employed to: 1) characterize changes in cervical length during pregnancy in a cohort of 5111 singleton pregnancies consisting predominantly of Black women; 2) test maternal risk factors for preterm birth, obstetric history and substance use domains as exogenous predictors of gestational age at delivery (GAD) and; 3) examine the extent to which cervical change parameters mediate associations between maternal risk factors and GAD. The underlying hypothesis was that changes in cervical length during pregnancy would, in part, mediate the effects of maternal risk factors, providing insight into etiologic mechanisms predisposing to preterm birth.

## METHODS

### Cohort Description

This study included women enrolled in a prospective cohort study of pregnant women at the Center for Advanced Obstetrical Care and Research (CAOCR) at Hutzel Women’s Hospital. The center was affiliated with Wayne State University and the Detroit Medical Center and part of the Pregnancy Research Branch (formerly Perinatology Research Branch) of The *Eunice Kennedy Shriver* National Institute of Child Health and Human Development (National Institutes of Health, U.S. Department of Health and Human Services). Clinical data collection was approved by the Institutional Review Boards of Wayne State University (#110605MP2F) and NICHD/NIH/DHHS (OH97-CH-N067). All study participants were enrolled between 2005 and 2017 and provided written informed consent.

From an initial cohort of 8226 pregnancies available with serial cervical length measurements, 5111 pregnancies were selected based on the following criteria: a singleton pregnancy, at least 2 cervical length measurements performed between 8 and 40 weeks of gestation, and availability of relevant demographic and clinical characteristics including self-identification as Black/African American. Women with a medically induced preterm delivery for any reason, history of cervical trauma, any serious medical conditions, or treatment with progesterone or cerclage during the study pregnancy were excluded in order to estimate model parameters on naturally occurring cervical length measures.

Demographic characteristics, relevant medical history, and pregnancy outcome data were obtained for each participant via medical record abstraction. Cervical length was measured in millimeters (mm) using a transvaginal 12-3 MHz ultrasound endocavitary probe by shearwave elastography (85). Serial cervical length measurements were obtained between 8 and 40 weeks of gestation when patients were seen for prenatal care visits in the CAOCR clinic. GAD was measured from the first day of a woman’s last menstrual period and confirmed by ultrasound. The inter-observer correlation of transvaginal cervical length was estimated based on 182 instances when a clinical and a research ultrasound evaluation were conducted in the same day for the same patients. The inter-observer correlation was 77%, similar to 76% reported elsewhere (47).

### Analytic Strategy

All modeling and data analyses in the current study were conducted in M*plus* (Version 8.3 for *Linux*) (86) using robust maximum likelihood estimation, where path coefficients and standard errors were computed while accounting for the non-independence of observations due to complex sampling (*i*.*e*., multiple pregnancies for the same woman, which accounted for 13.6% of pregnancies (n = 693)).

### Modeling Cervical Change Across Pregnancy

Growth curve modeling (GCM) is a cross-disciplinary analytic approach that utilizes repeated phenotypic measures to estimate intraindividual trajectories of phenotypic change, and interindividual differences in the parameters defining those trajectories. Contemporary applications of GCM typically derive from one of two distinct, but closely related methodological frameworks. The first, multilevel modeling (MLM) (87) was explicitly developed to allow for the specification of fixed and random effects in linear regression models where nested data structures are observed. The second, structural equation modeling (SEM) (88), extends Sewall Wright’s method of path coefficients (89) to test hypothesized associations between manifest and latent variables. Within SEM, latent growth curve analysis (LGCA) (90–92) characterizes the defining features of phenotypic trajectories via multiple indicator latent growth parameters. Given adequate sample size, a sufficient number of observations for each repeated measurement, and independence among study participants, univariate GCM conducted in MLM and SEM can be specified to yield equivalent estimates of the relevant parameters for all linear, and many nonlinear trajectories, including those conditioned on time-invariant and/or time-varying covariates (93).

Still, there are relevant advantages to selecting each framework over the other. For example, MLM is the preferable approach when the number and timing of repeated measures is highly variable across individuals (94). Conversely, SEM offers greater flexibility for testing various hypotheses (e.g., homoscedasticity of residual variances) (94), yields indices with which to formally evaluate model fit (95), and is more suitable for the analysis of complex mediation mechanisms (93). Moreover, since growth parameters in SEM are modeled as latent variables, and thusly disaggregated from measurement error, the approach is more psychometrically appealing (93). For these reasons, the method selected to analyze changes in cervical length across pregnancy was LGCA.

All LGCA were predicated on nine possible assessments of cervical length, each spanning a four-week window of observation with respect to the estimated date of conception (i.e., M2: 5-8 weeks; M3: 9-12 weeks; M4: 13-16 weeks; M5:17-20 weeks; M6: 21-24 weeks; M7: 25-28 weeks; M8: 29-32 weeks; M9: 33-36 weeks; M10: 37-40 weeks). For participants with two or more assessments of cervical length within a given four-week window, the mean of those values was used at that timepoint. Binning within temporal windows was necessary given sampling variability in the number and timing of individual observations distributed across 225 discrete days and importantly, produced parameter estimates that were comparable to those of MLM (Supplemental Table 1). As LGCA assumes a mean trajectory of change within the sample, and individual differences expressed in terms of normal variability around the specified growth parameters defining the level and shape of that change (90), a series of three nested models were estimated to determine which best characterized change in cervical length across pregnancy. The first, an intercept only or *no change* model, included a single latent growth parameter (I) reflecting the estimated mean cervical length at a specified temporal reference point within the process. In the present study, the intercept was set at M5 (i.e., 17-20 weeks gestation) to allow for comparison with the “mid-trimester” transvaginal sonographic measurement commonly reported in the literature; that is, after reproducible measurement is consistently possible (96), but before intervention for a short cervix is typically initiated (28). The second, a *linear change* model, included an additional latent growth parameter (L) reflecting the average change in cervical length per four weeks of gestation. The last, a *nonlinear change* model, included the addition of a third, quadratic growth parameter (Q) reflecting the mean rate of acceleration per four-week window of gestation. Figure 1 depicts a fully parameterized nonlinear change model that can easily be modified to represent a more restrictive no change or linear change model simply by fixing the factor loadings, variances, and covariances of the latent growth factor(s) L and/or Q, respectively, to zero. Finally, to determine whether variance components for the nine indicators of cervical length should be equated, as in MLM, models with and without this equality constraint were compared.

**Figure 1.**
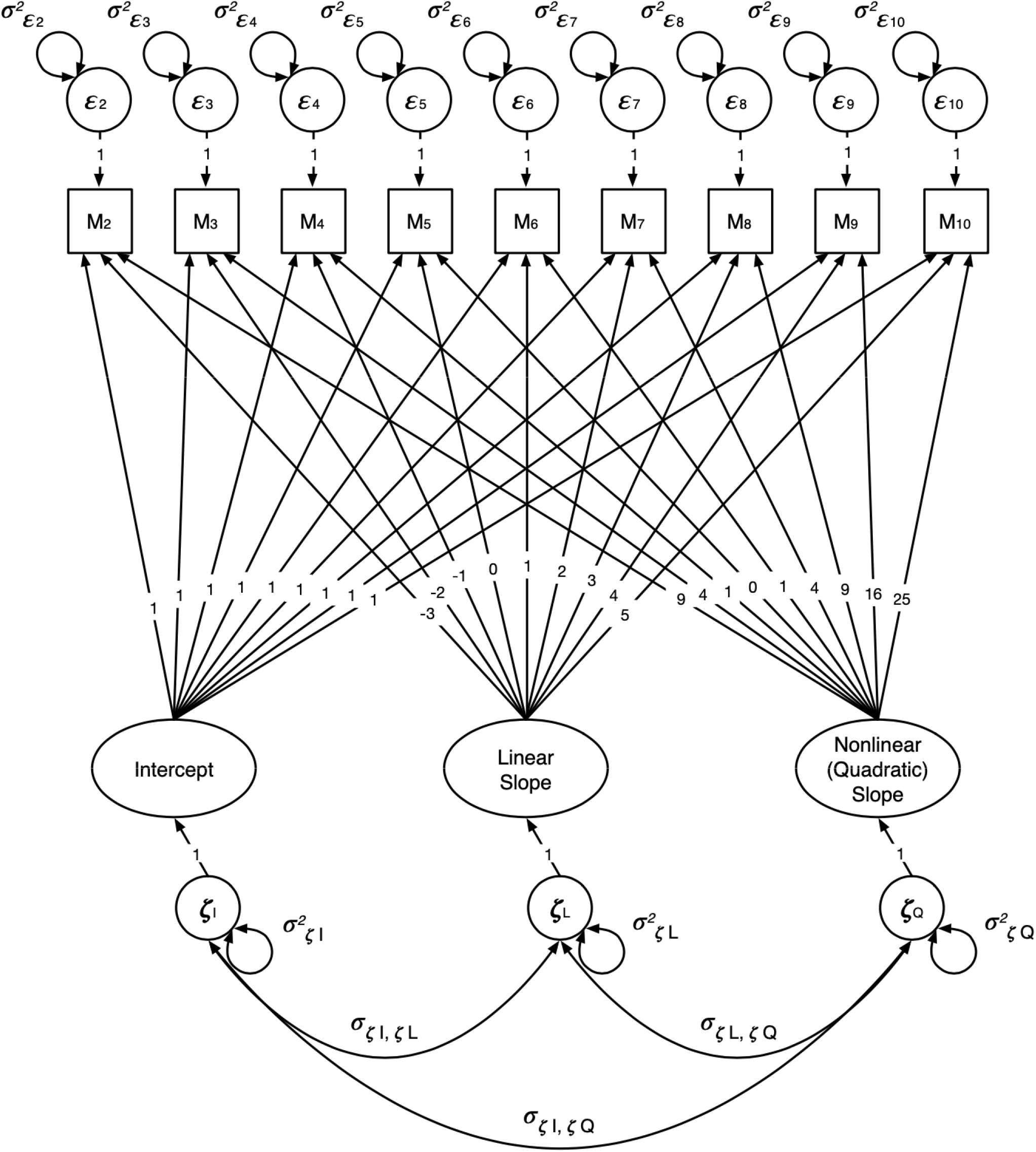
Parameterized latent growth curve analysis (LGCA) model. The intercept was set at M5 (i.e., month 5, 17-20 weeks gestation) to allow for comparison with the typical clinical assessment of the mid-trimester transvaginal sonographic measurement.

Interindividual differences in the number of cervical length assessments were expected for two reasons. First, measurements were obtained during regular prenatal care visits and without a prespecified schedule for the number of total visits and spacing between visits. Second, normal variability in the duration of gestation (i.e., term/preterm) was related to the participants’ time in the study, and thus the total number of recorded measurements. Among those pregnancies included in the unconditional LGCA, 344 of a possible 512 (i.e., 2^9^) data patterns were observed with respect to the nine monthly assessment periods. To summarize, 1.6% (n = 80) had representative data at all nine assessment periods, 63.0% (n = 3221) had representative data at four to eight assessment periods (i.e., surpassing the minimum threshold for non-linear analyses), and 35.4% (n = 1810) had representative data at one to three assessment periods.

### Multiple Mediation Modeling

A theoretically driven multiple mediation model was used to test the extent to which a set of known maternal risk factors accounted for variability in GAD, both directly and indirectly, through the latent growth factors described above, in order to characterize the intermediary role of changes in cervical length across the period of pregnancy. To avoid limitations of the traditional causal steps approach (97,98), a product of coefficients strategy (99,100) was implemented in M*plus* to disaggregate the total effect of an independent variable on a dependent variable (*c paths*) into direct (i.e., *X → Y*, or *c′ paths*) and indirect, or mediated effects (i.e., *X → M → Y*), all of which can be explicitly tested. In the case of simple mediation analysis, when an *X → Y* association is mediated through a single variable, the *indirect effect* is evaluated in relation to the *Z*-distribution, with the ratio of the product of the *a* (i.e., *X → M*) and *b* (i.e., *M → Y*) path coefficients over the normal-theory standard error for that product. Likewise, when an association is simultaneously mediated through multiple variables, the effect operating through a given variable (i.e., a *specific indirect effect*) is evaluated this same way. In contrast, when assessing the *total indirect effect* operating through multiple mediators, the sum of the products of the corresponding *a* and *b* path coefficients taken over the square root of the asymptotic variance of the sum of those products provides the ratio to be evaluated in relation to the *Z*-distribution.

## RESULTS

The filtered study cohort was comprised of 4,474 Black/African American women carrying 5,111 singleton pregnancies (Table 1). Of the 5,111 pregnancies in the cohort, 679 pregnancies (13.3%) were delivered preterm. Mid-trimester cervical length was measured between 18 and 24 weeks in 4,022 pregnancies (78.7%) and of these, 177 (4.40%) pregnancies met the clinical definition of a short cervix.

**Table 1.** Cohort Characteristics in a Population Sample of Pregnant Women from Detroit, Michigan, 2005-2013

| Variable | N | Overall, N = 4,824<sup>1</sup> | Preterm, N = 675 | Term, N = 4,149 | p-value<sup>2</sup> |
| --- | --- | --- | --- | --- | --- |
| **Maternal Age, Median (IQR)** | 4,824 | 23.0 (20.0 – 27.0) | 23.0 (20.0 – 28.0) | 23.0 (20.0 – 27.0) | 0.058 |
| **Depression, n (%)** | 4,824 | 140 (2.9) | 20 (3.0) | 120 (2.9) | 0.92 |
| **Pre-pregnancy BMI, Median (IQR)** | 4,824 | 28 (23 – 34) | 27 (22 – 34) | 28 (23 – 34) | 0.011 |
| **Parity, n (%)** | 4,022 |  |  |  | 0.12 |
| 0 |  | 1,701 (42) | 245 (46) | 1,456 (42) |  |
| 1 |  | 1,438 (36) | 172 (32) | 1,266 (36) |  |
| 2+ |  | 883 (22) | 113 (21) | 770 (22) |  |
| Unknown |  | 802 | 145 | 657 |  |
| **Previous Preterm Birth, n (%)** | 4,760 |  |  |  | <0.001 |
| 0 |  | 4,156 (87) | 472 (74) | 3,684 (89) |  |
| 1 |  | 504 (11) | 125 (20) | 379 (9.2) |  |
| 2+ |  | 100 (2.1) | 39 (6.1) | 61 (1.5) |  |
| Unknown |  | 64 | 39 | 25 |  |
| **Previous Abortion, n (%)** | 4,317 |  |  |  | 0.38 |
| 0 |  | 2,365 (55) | 315 (54) | 2,050 (55) |  |
| 1 |  | 1,310 (30) | 191 (33) | 1,119 (30) |  |
| 2+ |  | 642 (15) | 80 (14) | 562 (15) |  |
| Unknown |  | 507 | 89 | 418 |  |
| **Smoking, n (%)** | 4,810 | 922 (19) | 153 (23) | 769 (19) | 0.011 |
| Unknown |  | 14 | 2 | 12 |  |
| **Alcohol Use, n (%)** | 4,810 | 167 (3.5) | 26 (3.9) | 141 (3.4) | 0.55 |
| Unknown |  | 14 | 2 | 12 |  |
| **Substance Use, n (%)** | 4,815 | 1,383 (29) | 175 (26) | 1,208 (29) | 0.088 |
| Unknown |  | 9 | 1 | 8 |  |

<sup>1</sup> Median (IQR); n (%)
<sup>2</sup> Wilcoxon rank sum test; Pearson's Chi-squared test

### Characterizing Changes in Cervical Length Across Pregnancy

By way of LGCA, and standard practice for model selection within SEM (101), a combination of complementary fit indices corresponding to *no change, linear change*, and *nonlinear change* models of cervical length across pregnancy were compared (Table 2). The *χ*^*2*^ test of model fit for all three growth models exceeded significance thresholds for the specified degrees of freedom, yet this test alone was likely insufficient to suggest poor model fit, particularly in such a large sample. In contrast, standard thresholds for two sample size adjusted indices, the Comparative Fit Index (CFI > .95) (102) and the Root-Mean-Square Error of Approximation (RMSEA < .06) (101), suggested that the unconditional nonlinear model unequivocally provides the best representation of cervical change across pregnancy within this sample. Finally, to determine whether variance components across the nine indicators of cervical length should be equated, as in MLM, nonlinear growth models with and without this constraint were compared. A *χ*^*2*^ difference test scaled to accommodate non-normal data distributions (103) indicated that restricting the model in this way would lead to a significant decrement in model fit (*χ*^*2*^Δ_(4)_= 66.913, *p* < .001). As such, variances in the nine temporal indicators of cervical length were independently estimated.

**Table 2.** Model fit and growth parameter characteristics for unconditional latent growth curve models in a Population Sample of Pregnant Women from Detroit, Michigan, 2005-2013

|  | Model Fit |  |  |  |  | I |  | L |  | Q |  | Intercorrelations |  |  |
| --- | --- | --- | --- | --- | --- | --- | --- | --- | --- | --- | --- | --- | --- | --- |
| | $\chi^2$ | df | p-value | RMSEA | CFI | $\mu$ | $\sigma^2$ | $\mu$ | $\sigma^2$ | $\mu$ | $\sigma^2$ | $r_{IL}$ | $r_{LQ}$ | $r_{IQ}$ |
| No change | 4184.366 | 43 | <.0001 | .137 | .348 | 37.895 | 36.043 | -- | -- | -- | -- | -- | -- | -- |
| Linear change | 1274.635 | 40 | <.0001 | .078 | .806 | 38.928 | 25.091 | -1.047 | 1.697 | -- | -- | .539 | -- | -- |
| Nonlinear change | 165.232 | 36 | <.0001 | .027 | .980 | 39.813 | 28.245 | -.266 | 2.452 | -.333 | .081 | .664 | -.501 | -.530 |

*Note:* $\chi^2$ = chi-square test of model fit; RMSEA = root-mean-square error of approximation; CFI = comparative fit index; I = intercept (at 20-23 weeks gestation); L = linear slope; Q = quadratic slope (acceleration/deceleration); All growth parameter means, variances, and intercorrelations are significant at $p < .001$ .

A set of significant parameter estimates resulting from the nonlinear change model indicated that, on average: (1) cervical length during the fifth month of gestation (i.e., at the model intercept, from 17 through 20 weeks) was approximately 39.8 millimeters; (2) the change in cervical length across that same four-week period in gestation evidenced a modest decrease of just over one-quarter of a millimeter (*μ* _L_ = −.266 mm); and (3) this rate of cervical shortening accelerated by one-third of a millimeter per month post intercept (*μ* _Q_ = −.333), beginning at the vertex of the trajectory (i.e., at 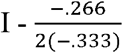 months), around the nineteenth week of pregnancy, prior to which a commensurate monthly deceleration was observed. Significant intraindividual variability was also observed in each of the growth parameters (all at *P* < .001), making it possible to include them as endogenous and/or exogenous components in larger structural models of association. Finally, significant intercorrelations among *I, L*, and *Q* suggest that women with longer cervical measurements at 17-20 weeks gestation display slower rates of cervical shortening at that point in gestation (*r*_IL_ = .664), but greater acceleration in that process across the remainder of the pregnancy (*r*_IQ_ = −.530). Likewise, women with flatter rates of cervical shortening at 17-20 weeks gestation are likely to experience more rapid decreases over the remaining weeks of pregnancy (*r*_LQ_ = −.501). For reference, the mean trajectory of accepted clinical preterm birth classes was plotted over a background of the raw cervical length data to demonstrate the difference in the overall functional forms of each trajectory compared to term births (Figure 2).

**Figure 2.**
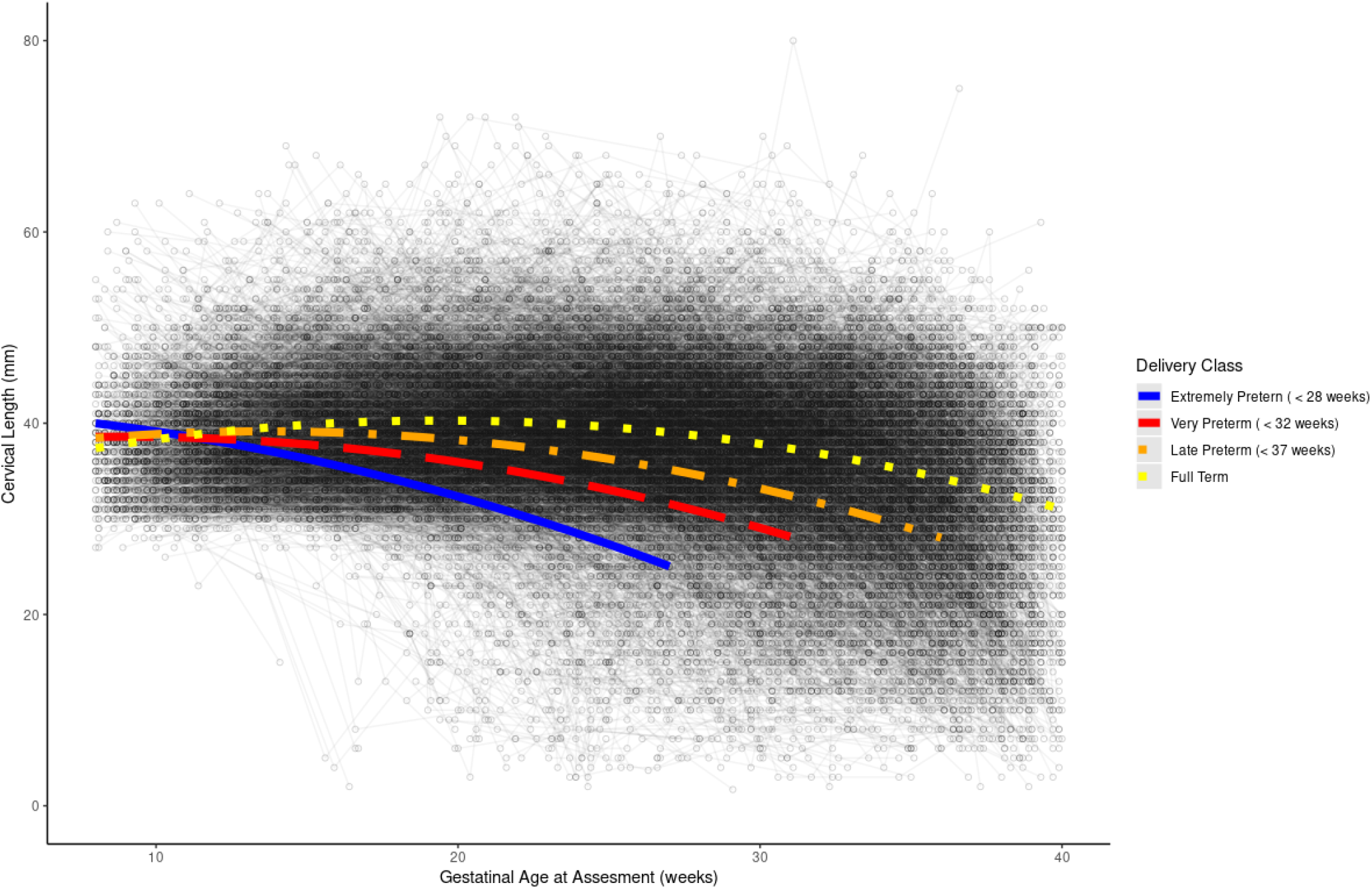
Longitudinal trajectories of cervical change by birth outcome class.

### Maternal Risk Factors Influencing GAD via Changes in Cervical Length

The nonlinear model of change in cervical length across pregnancy was incorporated into a larger SEM aimed at evaluating the influence of various maternal risk factors on GAD, both directly and indirectly, through intermediary influences on the latent factors characterizing cervical change: *I, L*, and *Q*. Representative variables from each of three broad domains of maternal characteristics commonly associated with risk for preterm birth were included as nine exogenous predictors of GAD: maternal health (*maternal age, maternal depression, pre-pregnancy BMI*), obstetric history (*parity, history of preterm births, history of spontaneous and induced abortions*), and substance use (*alcohol use, smoking*, and *other substance use*). Beyond regressing GAD onto the nine manifest indicators of maternal risk, it was additionally regressed onto *I, L*, and *Q*, which were, in turn, all concurrently regressed onto the maternal risk indicators. Figure 3 depicts the full, simultaneously estimated SEM, complete with standardized conditional path coefficients for all the bivariate associations described above (i.e., *c′, a*, and *b* paths). Overall, this model accounts for more than one-quarter of the sample variability in *GAD* (*R*^*2*^ = .265; *χ*^*2*^_(96)_ = 265.147, *p* < .001; RMSEA = .019; CFI = .980).

**Figure 3.**
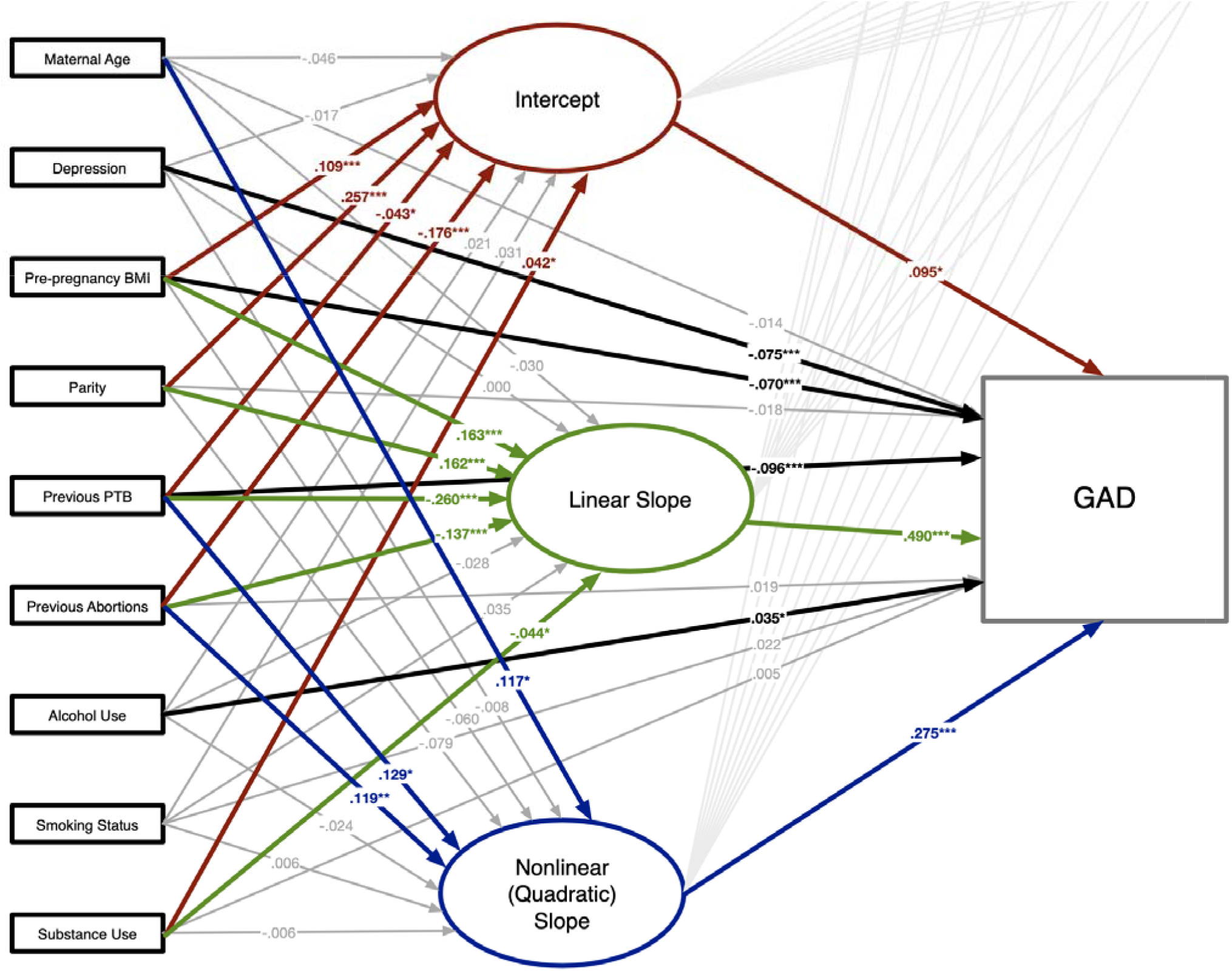
Full conditional mediation model with path coefficients. The total effects of individual maternal health, obstetric history, and substance use variables on gestational age at delivery (GAD) are unconditional with respect to the *I, L*, and *Q* factors characterizing cervical change, and not, therefore, explicitly depicted.

Table 3 provides a summary of the total, direct, and indirect effects for each of the nine manifest indicators of maternal risk in relation to GAD. Independent of cervical change, four of nine maternal risk variables have overall (total) effects on GAD, with maternal depression (*B*_*c*_ = −.062, *Z* = −3.128, *p* = 0.002) and previous PTB (*B*_*c*_ = −.204, *Z* = −10.416, *p* < 0.001) both predictive of a shorter gestational period, and parity (*B*_*c*_ = .064, *Z* = 3.480, *p* = 0.001) and smoking status (though somewhat less intuitive; *B*_*c*_ = .043, *Z* = 2.761, *p* = 0.006) both associated with a longer period of gestation. Although the total effect reflects the absolute influence of an exogenous predictor on an outcome conditional on other predictors, it can be parsed into the influence which is directly attributable to the predictor (a direct effect), and the influence of the predictor that is exerted indirectly, through one or more relevant ‘mediating’ variables (indirect effects), such that the total effect is equal to the sum of the direct and indirect effects. In the present model, only maternal age was shown to have neither direct nor indirect effects on GAD.

**Table 3.** Conditional mediated effects of maternal risk factors on gestational age at delivery via cervical change in a Population Sample of Pregnant Women from Detroit, Michigan, 2005-2013

| Maternal Risk | Direct Effect |  | Indirect Effect(s) |  | Total Effect |  |
| --- | --- | --- | --- | --- | --- | --- |
| | $B_{c'}(SE_{c'})$ | Ratio | $B_{a*b}(SE_{a*b})$ | Ratio | $B_c(SE_c)$ | Ratio |
| *Maternal Age* | -.014 (.012) | -1.175 | -.004 (.009) | -.420 | -.018 (.012) | -1.475 |
| specific IE via I ($a_I*b_I$) | | | -.002 (.002) | -1.030 | | |
| specific IE via L ($a_L*b_L$) | | | .000 (.010) | -.007 | | |
| specific IE via Q ($a_Q*b_Q$) | | | -.002 (.008) | -.246 | | |
| *Maternal Depression* | **-.075 (.020)** | **-3.727** | .013 (.014) | .975 | **-.062 (.020)** | **-3.128** |
| specific IE via I ($a_I*b_I$) | | | -.004 (.003) | -1.441 | | |
| specific IE via L ($a_L*b_L$) | | | -.014 (.015) | -.950 | | |
| specific IE via Q ($a_Q*b_Q$) | | | **.032 (.015)** | **2.132** | | |
| *Pre-pregnancy BMI* | **-.070 (.015)** | **-4.540** | **.074 (.010)** | **7.250** | .004 (.014) | .281 |
| specific IE via I ($a_I*b_I$) | | | **.010 (.005)** | **2.072** | | |
| specific IE via L ($a_L*b_L$) | | | **.080 (.013)** | **5.962** | | |
| specific IE via Q ($a_Q*b_Q$) | | | -.017 (.011) | -1.559 | | |
| *Parity* | -.018 (.019) | -.970 | **.082 (.013)** | **6.076** | **.064 (.018)** | **3.480** |
| specific IE via I ($a_I*b_I$) | | | **.024 (.011)** | **2.179** | | |
| specific IE via L ($a_L*b_L$) | | | **.079 (.017)** | **4.737** | | |
| specific IE via Q ($a_Q*b_Q$) | | | -.022 (.013) | -1.609 | | |
| *Previous PTB* | **-.096 (.021)** | **-4.586** | **-.108 (.016)** | **-6.980** | **-.204 (.020)** | **-10.416** |
| specific IE via I ($a_I*b_I$) | | | **-.017 (.008)** | **-2.135** | | |
| specific IE via L ($a_L*b_L$) | | | **-.127 (.020)** | **-6.241** | | |
| specific IE via Q ($a_Q*b_Q$) | | | **.036 (.016)** | **2.262** | | |
| *Previous Abortions* | .019 (.017) | 1.147 | **-.039 (.013)** | **-2.889** | -.020 (.016) | -1.214 |
| specific IE via I ($a_I*b_I$) | | | -.004 (.003) | -1.539 | | |
| specific IE via L ($a_L*b_L$) | | | **-.067 (.015)** | **-4.533** | | |
| specific IE via Q ($a_Q*b_Q$) | | | **.033 (.013)** | **2.517** | | |
| *Alcohol Use* | **.035 (.016)** | **2.121** | -.018 (.010) | -1.896 | .017 (.018) | .923 |
| specific IE via I ($a_I*b_I$) | | | .002 (.002) | .983 | | |
| specific IE via L ($a_L*b_L$) | | | -.013 (.010) | -1.335 | | |
| specific IE via Q ($a_Q*b_Q$) | | | -.007 (.010) | -.662 | | |
| *Smoking Status* | .022 (.015) | 1.415 | **.022 (.010)** | **2.200** | **.043 (.016)** | **2.761** |
| specific IE via I ($a_I*b_I$) | | | .003 (.002) | 1.326 | | |
| specific IE via L ($a_L*b_L$) | | | .017 (.011) | 1.503 | | |
| specific IE via Q ($a_Q*b_Q$) | | | .002 (.010) | .155 | | |
| *Substance Use* | .005 (.014) | .340 | **-.019 (.009)** | **-2.021** | -.014 (.014) | -1.000 |
| specific IE via I ($a_I*b_I$) | | | .004 (.002) | 1.660 | | |
| specific IE via L ($a_L*b_L$) | | | **-.021 (.011)** | **-1.977** | | |
| specific IE via Q ($a_Q*b_Q$) | | | -.002 (.010) | -.182 | | |

*Note:* All standardized effect coefficients and associated standard errors are rounded to the nearest thousandth, with significance depicted in boldface type. Critical ratios (i.e., *Z-scores*) of 1.96, 2.58, and 3.29 correspond to *p*-values of .05, .01, and .001, respectively. Total indirect effects (IE) for each maternal risk factor (i.e., $B_{a*b}$ ) disaggregate into three specific indirect effects, each operating through a single latent growth factor: the intercept (I), linear rate of change (L), or acceleration/deceleration (Q).

In terms of the four maternal risk factors with significant total effects, two retained direct effects on GAD in the presence of cervical change, with the influence of maternal depression slightly increasing in magnitude (*B*_*c′*_ = −.075, *Z* = −3.727, *p* < 0.001), and that of previous PTB decreasing by more than half (*B*_*c′*_ = -.096, *Z* = −4.586, *p* < 0.001). Although there was no significant overall indirect effect of maternal depression on GAD through the combination of *I, L*, and *Q*, there was one specific pathway through which GAD was modestly altered (*B*_*a*b* (Q)_ = .032, *Z* = 2.132, *p* = 0.033), that being the rate of acceleration in cervical change in the latter stages of gestation. This was offset by even smaller, nonsignificant changes in *I* and *L*. The effect of previous PTB on GAD was also mediated via changes in cervical length, as evidenced by an overall indirect effect (*B*_*a*b* Total_ = − 108, *Z* = −6.980, *p* < 0.001) that could be further disaggregated into three specific aspects of cervical change through which previous PTB significantly influenced the length of gestation (*B*_*a*b* (I)_ = −.017, *Z* = −2.135, *p* = 0.033; *B*_*a*b* (L)_ = − .127, *Z* = −6.241, *p* < 0.001; *B*_*a*b* (Q)_ = .036, *Z* = 2.262, *p* = 0.024). In contrast, the total effects of parity and smoking appear to be wholly attributable to changes in cervical length, as direct effects for both were nonsignificant. The indirect influence of parity via cervical change (*B*_*a*b* Total_ = .082, *Z* = 6.076, *p* < 0.001) was largely a function of variability in the length of the cervix and the expected rate of change at the 20-23 weeks gestation (*B*_*a*b* (I)_ = .024, *Z* = 2.179, *p* = 0.029; *B*_*a*b* (L)_ = .079, *Z* = 4.737, *p* < 0.001). The effect of maternal smoking status on GAD was best explained as operating indirectly through the overall process of cervical change (*B*_*a*b* Total_ = .022, *Z* = 2.200, *p* = 0.028), and not explicitly via any specific characteristic defining that process.

Four remaining indicators of maternal risk, none of which exhibited conditional overall (total) effects on GAD, did show direct and/or indirect effects when accounting for changes in the cervix. Specifically, direct effects emerged for both pre-pregnancy BMI (*B*_*c′*_ = −.070, *Z* = -4.540, *p* < 0.001) and alcohol use (*B*_*c′*_ =.035, *Z* = 2.121, *p* = 0.034) in the presence of cervical change indicators, suggesting negative and positive associations with GAD, respectively. The overall indirect effect of pre-pregnancy BMI (*B*_*a*b* Total_= .074, *Z* = 7.250, *p* < 0.001) via cervical change was largely driven by cervical length and expected rate of change in cervical length at 20-23 weeks gestation (*B*_*a*b* (I)_ = .010, *Z* = 2.072, *p* = 0.038; *B*_*a*b* (L)_ = .080, *Z* = 5.962, *p* < 0.001). The emergent direct effect of alcohol use on GAD can be explained, in part, by a near significant overall indirect effect via general changes in the cervix across pregnancy (*B*_*a*b* Total_= −.018, *Z* = 1.896, *p* = 0.058), though not attributable to any specific aspects of that process. Although neither total effects nor direct effects were observed in relation to previous abortions (spontaneous and induced) and substance use, both risk factors were shown to exert influence on GAD indirectly, through cervical change.

## DISCUSSION

In this study, longitudinal measurements of cervical length provided a unique opportunity to examine how changes in cervical length across pregnancy were related to common maternal risk factors of preterm birth. Multiple cervical length measures across pregnancy were summarized using LGCA methods to derive linear and non-linear indices of growth revealing unique associations with GAD. Mediational analyses provided insight on whether cervical length growth parameters accounted for the effect of maternal risk factors, not yet seen before in birth outcomes research. All analyses were performed on a large sample of Black women who historically account for the highest rate of health disparities in preterm birth and perinatal outcomes in the United States (104,105), have on average shorter mid-trimester cervical lengths than other racial/ethnic groups (53,83,106–108), and are represented by a disproportionately high burden of maternal risk factors (8,53,109,110).

As expected, the LGCA model including the nonlinear term provided the best fit to the data. Cervical length growth patterns by the specified preterm birth classes displayed a more rapid decrease in cervical length during pregnancy, while term births were associated with a gradual increase in cervical length before 25 weeks, followed by a gradual decrease in cervical length until delivery. Note that this may not reflect a physiological “lengthening” of the cervix, but rather changes in the structure, such as increased hydration or swelling, that produce slightly longer cervical length measurements than earlier in pregnancy (60). The mean estimate for cervical length at each timepoint also begins to differ significantly between preterm birth classes (see Figure 2), around 16 weeks of gestation, which is consistent with the literature for using mid-trimester cervical length between 18 and 24 weeks to predict preterm birth (28).

The growth model intercept term was parameterized to correspond to the mean mid-trimester cervical length, which is utilized clinically as a diagnostic criterion, along with other maternal risk factors, to identify women at elevated preterm birth risk. These results demonstrated that other aspects of the cervix during pregnancy can provide insight into the relationship between maternal risk factors and birth outcomes. In fact, none of the maternal risk factors were mediated exclusively through the intercept term, while maternal depression, substance use, and previous abortion were mediated via either the linear and/or non-linear terms. These results suggest that cervical change across pregnancy was more informative than a single mid-trimester cervical length measurement as a mediator of the relationship between maternal characteristics and PTB risk.

For example, the total effect of parity was found to be fully mediated through the intercept and linear change terms of the cervical length growth model. This result is consistent with the finding that parity is associated with preterm birth (49,50,111) and provides context for future investigations. For instance, the proper functioning of the cervix and mechanical support of the developing fetus early in pregnancy might function as a rate limiting step as reflected in the importance of the growth model terms that corresponded to the overall length of the cervix (*I*) and earlier cervical change (*S*). This contrasted with the finding that a previous preterm birth (spontaneous or induced) was associated with lower GAD and was only partially mediated through cervical change, albeit through all growth parameter terms. This finding could imply that the effect of a previous early birth on GAD is not only mediated by cervical dysfunction (e.g., cervical tissue damage) but may involve other pathways not related to cervical function (e.g., vascular, hormonal, psychological stress pathways).

One potential study limitation was that the number and spacing of participant visits were influenced by the prenatal care needs specific to each pregnancy. This lack of standardization in scheduling may have affected study results in unknown ways. Not surprisingly, the number of participant observations was associated with error variance in LGCA model parameters. Yet, only minimal differences in mean error variance for I, S, and Q were observed, for instance, between term and preterm pregnancies (I: 9.22 vs 9.39; S: 2.96 vs 3.12; Q: 0.047 vs 0.054). Parameterization of the LGCA model was predicated on the binning of cervical measurements into nine 4-week intervals which would necessarily exclude the last bin (37-40 weeks), at minimum, from preterm births. Although the absence of observations for preterm participants in the latter bins would not technically be considered missing, as they were not expected, parameter estimates from LGCA models were still comparable to those from MLM which did not require binning, as described previously (Supplemental Table 1). In addition, preterm birth status was not associated with the absence of a cervical length measurement when a study visit could be expected (Supplemental Figure 1). Another limitation was the racial composition of the study cohort, comprised of women who self-identified as Black/African American, and although the findings of this study may not be generalizable to women from other populations or ancestry groups, they could improve screening and clinical care for a population of women who are disproportionally affected by health disparities in preterm birth and other perinatal outcomes. Finally, mediation models can be conceptually used to assess potential causal mechanisms yet by themselves do not prove the proposed causal pathway.

The unique data source used, reflecting several repeated measures across pregnancy, provided a rare opportunity to identify etiologic pathways for the influence of preterm birth risk factors operating, at least in part, through cervical length changes during pregnancy.

## Data Availability

All data produced in the present study are available upon reasonable request to the authors.

## ETHICS APPROVAL

The Institutional Review Boards of Wayne State University and the Eunice Kennedy Shriver National Institute of Child Health and Human Development (NICHD)/National Institutes of Health/U.S. Department of Health and Human Services (Detroit, MI, USA) approved the study. Participants were enrolled under the protocols Biological Markers of Disease in the Prediction of Preterm Delivery, Preeclampsia and Intra-Uterine Growth Restriction: A Longitudinal Study (WSU IRB#110605MP2F and NICHD/NIH# OH97-CH-N067). All participants provided written informed consent for the collection of cervical length data and blood samples for future genetic research studies.

## SUPPLEMENTAL MATERIALS

**Table S1.** Comparison of parameter estimates from Latent Growth Curve Analysis and Multilevel Models.

| Modelling Method | I Estimate<br>Median (Range) | L Estimate<br>Median (Range) | Q Estimate<br>Median (Range) |
| --- | --- | --- | --- |
| *Multilevel Model (R {lmer})*<br>Non-binned data | 35.9<br>(28.5, 43.9) | 2.23<br>(-5.5, 8.8) | -0.35<br>(-1.09, 0.49) |
| *Multilevel Model (R {nlme})*<br>Binned data | 37.0<br>(31.8, 49.0) | 1.87<br>(-9.05, 11.1) | -0.32<br>(-1.11, 0.79) |
| *Multilevel Model (Mplus)*<br>Binned data | 37.1<br>(31.2, 49.1) | 1.88<br>(-10.3, 10.9) | -0.33<br>(-1.09, 0.57) |
| *Latent Growth Curve (Mplus)*<br>Binned data | 38.5<br>(30.9, 46.8) | 1.30<br>(-9.98, 8.14) | -0.34<br>(-1.02, 0.66) |

**Figure S1.**
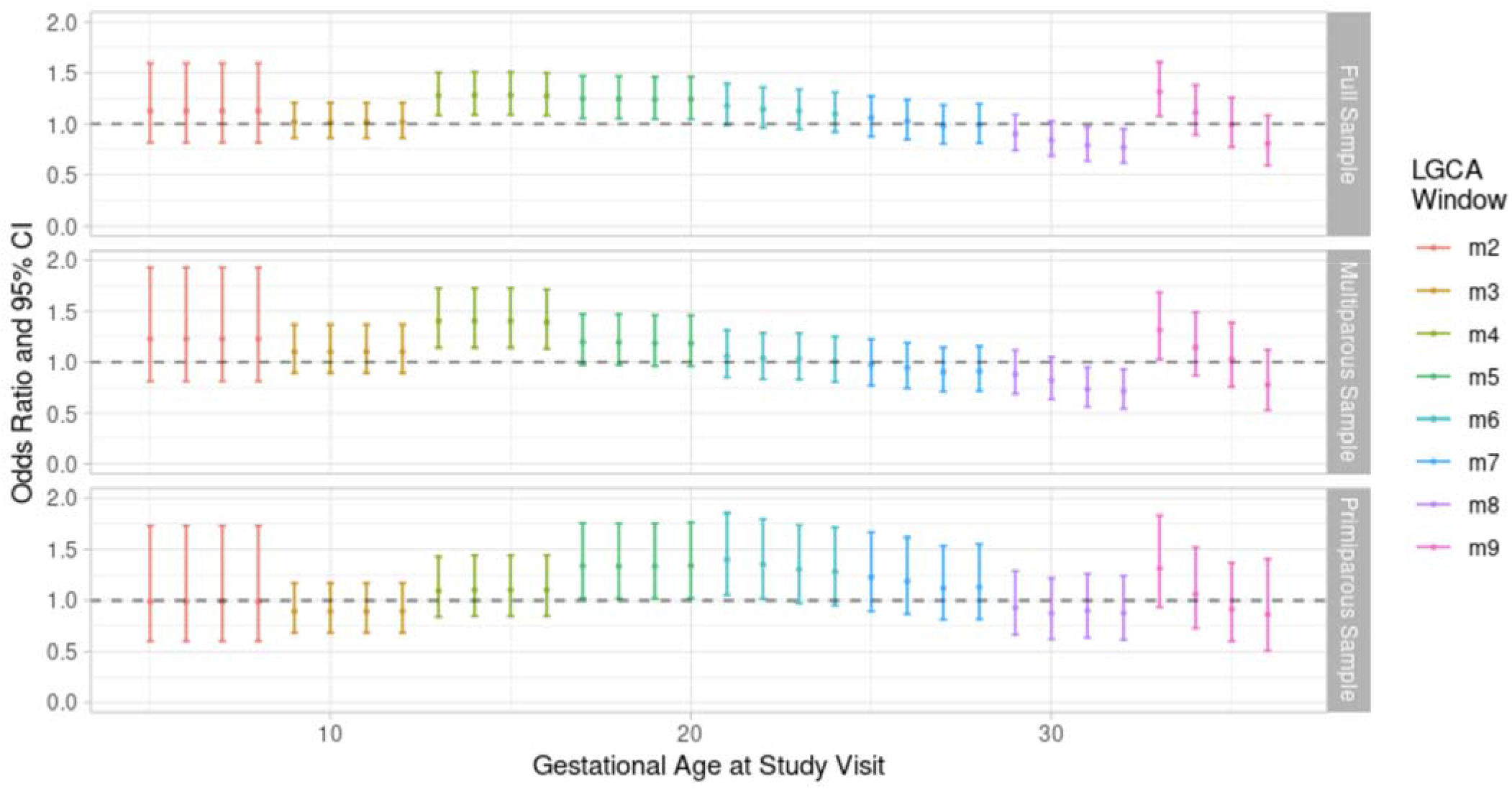
Relationship between the index pregnancy being delivered preterm and presence of cervical length measure at each gestational age. Increased odds ratios observed in M4 can be explained by parity status which was controlled for in all mediation models.

## REFERENCES

1. Blencowe H, Cousens S, Chou D, et al. Born Too Soon: The global epidemiology of 15 million preterm births. Reprod Health. 2013;10(Suppl 1):S2.

2. Althabe F, Howson CP, Kinney M, et al. Born too soon: the global action report on preterm birth. 2012.

3. Andersen HF, Nugent CE, Wanty SD, et al. Prediction of risk for preterm delivery by ultrasonographic measurement of cervical length. American Journal of Obstetrics and Gynecology. 1990;163(3):859–867.

4. Iams JD, Goldenberg RL, Meis PJ, et al. The Length of the Cervix and the Risk of Spontaneous Premature Delivery. New England Journal of Medicine. 1996;334(9):567–573.

5. Berghella V, Tolosa JE, Kuhlman K, et al. Cervical ultrasonography compared with manual examination as a predictor of preterm delivery. American Journal of Obstetrics and Gynecology. 1997;177(4):723–730.

6. Goldenberg RL, Iams JD, Mercer BM, et al. The preterm prediction study: the value of new vs standard risk factors in predicting early and all spontaneous preterm births. NICHD MFMU Network. Am J Public Health. 1998;88(2):233–238.

7. Guzman ER, Mellon C, Vintzileos AM, et al. Longitudinal assessment of endocervical canal length between 15 and 24 weeks’ gestation in women at risk for pregnancy loss or preterm birth. Obstet Gynecol. 1998;92(1):31–37.

8. Heath VC, Southall TR, Souka AP, et al. Cervical length at 23 weeks of gestation: relation to demographic characteristics and previous obstetric history. Ultrasound Obstet Gynecol. 1998;12(5):304–311.

9. Hassan SS, Romero R, Berry SM, et al. Patients with an ultrasonographic cervical length ≤15 mm have nearly a 50% risk of early spontaneous preterm delivery. American Journal of Obstetrics & Gynecology. 2000;182(6):1458–1467.

10. Hibbard JU, Tart M, Moawad AH. Cervical length at 16–22 weeks’ gestation and risk for preterm delivery. Obstetrics & Gynecology. 2000;96(6):972–978.

11. Owen J, Yost N, Berghella V, et al. Mid-Trimester Endovaginal Sonography in Women at High Risk for Spontaneous Preterm Birth. JAMA. 2001;286(11):1340–1348.

12. Honest H, Bachmann LM, Coomarasamy A, et al. Accuracy of cervical transvaginal sonography in predicting preterm birth: a systematic review. Ultrasound in Obstetrics & Gynecology. 2003;22(3):305–322.

13. Owen J, Yost N, Berghella V, et al. Can shortened midtrimester cervical length predict very early spontaneous preterm birth? American Journal of Obstetrics and Gynecology. 2004;191(1):298–303.

14. To MS, Skentou CA, Royston P, et al. Prediction of patient-specific risk of early preterm delivery using maternal history and sonographic measurement of cervical length: a population-based prospective study. Ultrasound in Obstetrics & Gynecology. 2006;27(4):362–367.

15. Crane JMG, Hutchens D. Transvaginal sonographic measurement of cervical length to predict preterm birth in asymptomatic women at increased risk: a systematic review. Ultrasound in Obstetrics & Gynecology. 2008;31(5):579–587.

16. Domin CM, Smith EJ, Terplan M. Transvaginal ultrasonographic measurement of cervical length as a predictor of preterm birth: a systematic review with meta-analysis. Ultrasound Q. 2010;26(4):241–248.

17. Vaisbuch E, Romero R, Erez O, et al. Clinical significance of early (< 20 weeks) vs. late (20–24 weeks) detection of sonographic short cervix in asymptomatic women in the mid-trimester. Ultrasound in Obstetrics & Gynecology. 2010;36(4):471–481.

18. Berghella V. Universal cervical length screening for prediction and prevention of preterm birth. Obstet Gynecol Surv. 2012;67(10):653–658.

19. da Fonseca EB, Bittar RE, Carvalho MHB, et al. Prophylactic administration of progesterone by vaginal suppository to reduce the incidence of spontaneous preterm birth in women at increased risk: A randomized placebo-controlled double-blind study. American Journal of Obstetrics and Gynecology. 2003;188(2):419–424.

20. Fonseca EB, Celik E, Parra M, et al. Progesterone and the risk of preterm birth among women with a short cervix. N. Engl. J. Med. 2007;357(5):462–469.

21. Cetingoz E, Cam C, Sakalli M, et al. Progesterone effects on preterm birth in high-risk pregnancies: a randomized placebo-controlled trial. Arch Gynecol Obstet. 2011;283(3):423–429.

22. Hassan SS, Romero R, Vidyadhari D, et al. Vaginal progesterone reduces the rate of preterm birth in women with a sonographic short cervix: a multicenter, randomized, double-blind, placebo-controlled trial. Ultrasound in Obstetrics & Gynecology. 2011;38(1):18–31.

23. Combs CA. Vaginal progesterone for asymptomatic cervical shortening and the case for universal screening of cervical length. American Journal of Obstetrics and Gynecology. 2012;206(2):101–103.

24. Romero R, Nicolaides K, Conde-Agudelo A, et al. Vaginal progesterone in women with an asymptomatic sonographic short cervix in the midtrimester decreases preterm delivery and neonatal morbidity: a systematic review and metaanalysis of individual patient data. American Journal of Obstetrics and Gynecology. 2012;206(2):124.e1-124.e19.

25. Practice Bulletin No. 130: Prediction and Prevention of Preterm Birth. Obstetrics & Gynecology. 2012;120(4):964–973.

26. Society for Maternal-Fetal Medicine Publications Committee. Progesterone and preterm birth prevention: translating clinical trials data into clinical practice. American Journal of Obstetrics and Gynecology. 2012;206(5):376–386.

27. Conde-Agudelo A, Romero R, Nicolaides K, et al. Vaginal progesterone vs cervical cerclage for the prevention of preterm birth in women with a sonographic short cervix, previous preterm birth, and singleton gestation: a systematic review and indirect comparison metaanalysis. American Journal of Obstetrics and Gynecology. 2013;208(1):42.e1-42.e18.

28. Romero R, Yeo L, Miranda J, et al. A blueprint for the prevention of preterm birth: vaginal progesterone in women with a short cervix. Journal of Perinatal Medicine. 2013;41(1):27–44.

29. Furcron A-E, Romero R, Plazyo O, et al. Vaginal progesterone, but not 17α-hydroxyprogesterone caproate, has antiinflammatory effects at the murine maternal-fetal interface. American Journal of Obstetrics and Gynecology. 2015;213(6):846.e1-846.e19.

30. Conde-Agudelo A, Romero R. Vaginal progesterone to prevent preterm birth in pregnant women with a sonographic short cervix: clinical and public health implications. American Journal of Obstetrics and Gynecology. 2016;214(2):235–242.

31. Vintzileos AM, Visser GHA. Interventions for women with mid-trimester short cervix: which ones work?: Editorial. Ultrasound in Obstetrics & Gynecology. 2017;49(3):295–300.

32. Conde-Agudelo A, Romero R, Da Fonseca E, et al. Vaginal progesterone is as effective as cervical cerclage to prevent preterm birth in women with a singleton gestation, previous spontaneous preterm birth, and a short cervix: updated indirect comparison meta-analysis. American Journal of Obstetrics and Gynecology. 2018;219(1):10–25.

33. Romero R, Conde-Agudelo A, Da Fonseca E, et al. Vaginal progesterone for preventing preterm birth and adverse perinatal outcomes in singleton gestations with a short cervix: a meta-analysis of individual patient data. American Journal of Obstetrics and Gynecology. 2018;218(2):161–180.

34. Berghella V, Rafael TJ, Szychowski JM, et al. Cerclage for Short Cervix on Ultrasonography in Women With Singleton Gestations and Previous Preterm Birth: A Meta-Analysis. Obstetrics & Gynecology. 2011;117(3):663–671.

35. Daskalakis G, Loutradis D, Antsaklis A, et al. A stepwise approach for the management of short cervix: time to evolve beyond progesterone treatment in the presence of progressive cervical shortening. Am J Obstet Gynecol. 2019;220(4):404–405.

36. Roman A, Zork N, Haeri S, et al. Physical examination-indicated cerclage in twin pregnancy: a randomized controlled trial. Am J Obstet Gynecol. 2020;223(6):902.e1-902.e11.

37. Romero R, Espinoza J, Erez O, et al. The role of cervical cerclage in obstetric practice: can the patient who could benefit from this procedure be identified? Am J Obstet Gynecol. 2006;194(1):1–9.

38. Conde-Agudelo A, Romero R. Does vaginal progesterone prevent recurrent preterm birth in women with a singleton gestation and a history of spontaneous preterm birth? Evidence from a systematic review and meta-analysis. Am J Obstet Gynecol. 2022;227(3):440-461.e2.

39. Conde-Agudelo A, Romero R. Vaginal progesterone does not prevent recurrent preterm birth in women with a singleton gestation, a history of spontaneous preterm birth, and a midtrimester cervical length >25 mm. Am J Obstet Gynecol. 2022;227(6):923–926.

40. Conde-Agudelo A, Romero R. Vaginal progesterone for the prevention of preterm birth: who can benefit and who cannot? Evidence-based recommendations for clinical use. J Perinat Med. 2023;51(1):125–134.

41. House M, Tadesse-Telila S, Norwitz ER, et al. Inhibitory Effect of Progesterone on Cervical Tissue Formation in a Three-Dimensional Culture System with Human Cervical Fibroblasts. Biol Reprod. 2014;90(1):18.

42. Conde-Agudelo A, Romero R. Predictive accuracy of changes in transvaginal sonographic cervical length over time for preterm birth: a systematic review and metaanalysis. American Journal of Obstetrics and Gynecology. 2015;213(6):789–801.

43. Wolf HM, Romero R, Strauss JF, et al. Study protocol to quantify the genetic architecture of sonographic cervical length and its relationship to spontaneous preterm birth. BMJ Open. 2022;12(3):e053631.

44. Word RA, Li X-H, Hnat M, et al. Dynamics of Cervical Remodeling during Pregnancy and Parturition: Mechanisms and Current Concepts. Semin Reprod Med. 2007;25(01):069–079.

45. House M, Kaplan DL, Socrate S. Relationships between Mechanical Properties and Extracellular Matrix Constituents of the Cervical Stroma during Pregnancy. Semin Perinatol. 2009;33(5):300–307.

46. Holt R, Timmons BC, Akgul Y, et al. The molecular mechanisms of cervical ripening differ between term and preterm birth. Endocrinology. 2011;152(3):1036–1046.

47. Gudicha DW, Romero R, Kabiri D, et al. Personalized assessment of cervical length improves prediction of spontaneous preterm birth: a standard and a percentile calculator. American Journal of Obstetrics and Gynecology. 2021;224(3):288.e1–288.e17.

48. Rosenbloom JI, Raghuraman N, Temming LA, et al. Predictive Value of Midtrimester Universal Cervical Length Screening Based on Parity. Journal of Ultrasound in Medicine. 2020;39(1):147–154.

49. Berkowitz GS, Blackmore-Prince C, Lapinski RH, et al. Risk Factors for Preterm Birth Subtypes. Epidemiology. 1998;9(3):279.

50. Ananth CV, Peltier MR, Getahun D, et al. Primiparity: an “intermediate” risk group for spontaneous and medically indicated preterm birth. J Matern Fetal Neonatal Med. 2007;20(8):605–611.

51. Andersen HF. Transvaginal and transabdominal ultrasonography of the uterine cervix during pregnancy. J Clin Ultrasound. 1991;19(2):77–83.

52. Petrović D, Novakov-Mikić A, Mandić V. Socio-demographic factors and cervical length in pregnancy. Med. Pregl. 2008;61(9–10):443–451.

53. van der Ven AJ, van Os MA, Kleinrouweler CE, et al. Is cervical length associated with maternal characteristics? European Journal of Obstetrics & Gynecology and Reproductive Biology. 2015;188:12–16.

54. Berghella V, Roman A, Daskalakis C, et al. Gestational age at cervical length measurement and incidence of preterm birth. Obstet Gynecol. 2007;110(2 Pt 1):311–317.

55. Timmons B, Akins M, Mahendroo M. Cervical Remodeling during Pregnancy and Parturition. Trends Endocrinol Metab. 2010;21(6):353–361.

56. Hassan SS, Romero R, Tarca AL, et al. The transcriptome of cervical ripening in human pregnancy before the onset of labor at term: identification of novel molecular functions involved in this process. J. Matern. Fetal. Neonatal. Med. 2009;22(12):1183–1193.

57. Hassan SS, Romero R, Tarca AL, et al. The molecular basis for sonographic cervical shortening at term: identification of differentially expressed genes and the epithelial-mesenchymal transition as a function of cervical length. American Journal of Obstetrics & Gynecology. 2010;203(5):472.e1-472.e14.

58. Timmons BC, Fairhurst A, Mahendroo MS. Temporal Changes in Myeloid Cells in the Cervix during Pregnancy and Parturition. J Immunol. 2009;182(5):2700–2707.

59. Read CP, Word RA, Ruscheinsky MA, et al. Cervical remodeling during pregnancy and parturition: molecular characterization of the softening phase in mice. Reproduction. 2007;134(2):327–340.

60. Myers KM, Hendon CP, Gan Y, et al. A continuous fiber distribution material model for human cervical tissue. Journal of Biomechanics. 2015;48(9):1533–1540.

61. Yoshida K, Jayyosi C, Lee N, et al. Mechanics of cervical remodelling: insights from rodent models of pregnancy. Interface Focus. 2019;9(5):20190026.

62. Timmons BC, Mitchell SM, Gilpin C, et al. Dynamic changes in the cervical epithelial tight junction complex and differentiation occur during cervical ripening and parturition. Endocrinology. 2007;148(3):1278–1287.

63. Parikh R, Patel A, Stack T, et al. How the cervix shortens: an anatomic study using 3-dimensional transperineal sonography and image registration in singletons and twins. J Ultrasound Med. 2011;30(9):1197–1204.

64. House M, McCabe R, Socrate S. Using imaging-based, three-dimensional models of the cervix and uterus for studies of cervical changes during pregnancy. Clinical Anatomy. 2013;26(1):97–104.

65. Myers KM, Feltovich H, Mazza E, et al. The mechanical role of the cervix in pregnancy. J Biomech. 2015;48(9):1511–1523.

66. Moroz LA, Simhan HN. Rate of sonographic cervical shortening and the risk of spontaneous preterm birth. American Journal of Obstetrics and Gynecology. 2012;206(3):234.e1-234.e5.

67. Salomon LJ, DiazLGarcia C, Bernard JP, et al. Reference range for cervical length throughout pregnancy: non-parametric LMS-based model applied to a large sample. Ultrasound in Obstetrics & Gynecology. 2009;33(4):459–464.

68. Bergelin I, Valentin L. Patterns of normal change in cervical length and width during pregnancy in nulliparous women: a prospective, longitudinal ultrasound study. Ultrasound Obstet Gynecol. 2001;18(3):217–222.

69. Bergelin I, Valentin L. Normal cervical changes in parous women during the second half of pregnancy – a prospective, longitudinal ultrasound study. Acta Obstetricia et Gynecologica Scandinavica. 2002;81(1):31–38.

70. Mercer BM, Goldenberg RL, Meis PJ, et al. The Preterm Prediction Study: Prediction of preterm premature rupture of membranes through clinical findings and ancillary testing. American Journal of Obstetrics & Gynecology. 2000;183(3):738–745.

71. Odibo AO, Berghella V, Reddy U, et al. Does transvaginal ultrasound of the cervix predict preterm premature rupture of membranes in a high-risk population? Ultrasound in Obstetrics & Gynecology. 2001;18(3):223–227.

72. Romero R, Gonzalez R, Sepulveda W, et al. Infection and labor: VIII. Microbial invasion of the amniotic cavity in patients with suspected cervical incompetence: Prevalence and clinical significance. American Journal of Obstetrics and Gynecology. 1992;167(4, Part 1):1086–1091.

73. Meis PJ, Goldenberg RL, Mercer B, et al. The preterm prediction study: Significance of vaginal infections. American Journal of Obstetrics and Gynecology. 1995;173(4):1231–1235.

74. Hassan SS, Romero R, Hendler I, et al. A sonographic short cervix as the only clinical manifestation of intra-amniotic infection. J Perinat Med. 2006;34(1):13–19.

75. Naim A, Haberman S, Burgess T, et al. Changes in cervical length and the risk of preterm labor. American Journal of Obstetrics and Gynecology. 2002;186(5):887–889.

76. Gomez R, Romero R, Nien JK, et al. A short cervix in women with preterm labor and intact membranes: A risk factor for microbial invasion of the amniotic cavity. American Journal of Obstetrics and Gynecology. 2005;192(3):678–689.

77. Sotiriadis A, Papatheodorou S, Kavvadias A, et al. Transvaginal cervical length measurement for prediction of preterm birth in women with threatened preterm labor: a meta-analysis. Ultrasound in Obstetrics & Gynecology. 2010;35(1):54–64.

78. Kandil M, Sanad Z, Sayyed T, et al. Body mass index is linked to cervical length and duration of pregnancy: An observational study in low risk pregnancy. J Obstet Gynaecol. 2017;37(1):33–37.

79. Igel C, Dar P, Rosner M, et al. High Maternal BMI Associated With Cervical Shortening in Women With Short Cervix on Second Trimester Anatomy Scan [27K]. Obstetrics & Gynecology. 2016;127:96S.

80. Buck JN, Orzechowski KM, Berghella V. Racial disparities in cervical length for prediction of preterm birth in a low risk population. The Journal of Maternal-Fetal & Neonatal Medicine. 2017;30(15):1851–1854.

81. Findley J, Seybold DJ, Broce M, et al. Transvaginal Cervical Length and Tobacco Use in Appalachian Women: Association with Increased Risk for Spontaneous Preterm Birth. W V Med J. 2015;111(3):22–28.

82. Cho S-H, Park KH, Jung EY, et al. Maternal Characteristics, Short Mid-Trimester Cervical Length, and Preterm Delivery. J Korean Med Sci. 2017;32(3):488–494.

83. Harville EW, Knoepp LR, Wallace ME, et al. Cervical pathways for racial disparities in preterm births: the Preterm Prediction Study. The Journal of Maternal-Fetal & Neonatal Medicine. 2018;0(0):1–7.

84. Brittain JJ, Wahl SE, Strauss JF, et al. Prior Spontaneous or Induced Abortion Is a Risk Factor for Cervical Dysfunction in Pregnant Women: a Systematic Review and Meta-analysis. Reprod Sci. 2023;

85. Berghella V, Bega G, Tolosa JE, et al. Ultrasound Assessment of the Cervix. Clinical Obstetrics and Gynecology. 2003;46(4):947.

86. Muthén LK, Muthén BO. Mplus User’s Guide. Eighth Edition. 2017;(https://www.statmodel.com/ugexcerpts.shtml). (Accessed October 7, 2019)

87. Goldstein H. Multilevel Models in Educational and Social Research. Oxford University Press, London: 1987.

88. Hoyle RH. The structural equation modeling approach: Basic concepts and fundamental issues. In: Structural equation modeling: Concepts, issues, and applications. Thousand Oaks, CA, US: Sage Publications, Inc; 1995:1–15.

89. Wright S. The method of path coefficients. Annals of Mathematical Statistics. 1934;5:161–215.

90. McArdle JJ, Epstein D. Latent growth curves within developmental structural equation models. Child Dev. 1987;58(1):110–133.

91. Meredith W, Tisak J. Latent curve analysis. Psychometrika. 1990;55:107–122.

92. Willett JB, Sayer AG. Using covariance structure analysis to detect correlates and predictors of individual change over time. Psychological Bulletin. 1994;116:363–381.

93. Curran PJ. Have Multilevel Models Been Structural Equation Models All Along? Multivariate Behav Res. 2003;38(4):529–569.

94. Stoel RD, van den Wittenboer G, Hox J. Analyzing Longitudinal Data using Multilevel Regression and Latent Growth Curve Analysis. Metodologia de las Ciencias del Comportamiento. 2003;5:21–42.

95. Chou C, Bentler PM, Pentz MA. Comparisons of two statistical approaches to study growth curves: The multilevel model and the latent curve analysis. Structural Equation Modeling: A Multidisciplinary Journal. 1998;5(3):247–266.

96. Berghella V, Talucci M, Desai A. Does transvaginal sonographic measurement of cervical length before 14 weeks predict preterm delivery in highLrisk pregnancies? Ultrasound in Obstetrics & Gynecology. 2003;21(2):140–144.

97. Baron RM, Kenny DA. The moderator-mediator variable distinction in social psychological research: conceptual, strategic, and statistical considerations. Journal of personality and social psychology. 1986;51(6):1173–1182.

98. Judd CM, Kenny DA. Process Analysis: Estimating Mediation in Treatment Evaluations. Eval Rev. 1981;5(5):602–619.

99. MacKinnon DP, Lockwood CM, Hoffman JM, et al. A comparison of methods to test mediation and other intervening variable effects. Psychol Methods. 2002;7(1):83–104.

100. Preacher KJ, Hayes AF. SPSS and SAS procedures for estimating indirect effects in simple mediation models. Behavior Research Methods, Instruments & Computers. 2004;36:717–731.

101. Hu L, Bentler PM. Cutoff criteria for fit indexes in covariance structure analysis: Conventional criteria versus new alternatives. Structural Equation Modeling: A Multidisciplinary Journal. 1999;6(1):1–55.

102. Bentler PM. Comparative fit indexes in structural models. Psychological Bulletin. 1990;107:238–246.

103. Satorra A, Bentler PM. Ensuring Positiveness of the Scaled Difference Chi-square Test Statistic. Psychometrika. 2010;75(2):243–248.

104. Culhane JF, Goldenberg RL. Racial Disparities in Preterm Birth. Seminars in Perinatology. 2011;35(4):234–239.

105. Spriggs AL. Racial disparities in preterm birth: the role of social determinants. American Journal of Obstetrics and Gynecology. 2007;197(3):328.

106. Dijkstra K, Janssen HCJP, Kuczynski E, et al. Cervical length in uncomplicated pregnancy: A study of sociodemographic predictors of cervical changes across gestation. American Journal of Obstetrics and Gynecology. 1999;180(3):639–644.

107. Harville EW, Miller KS, Knoepp LR. Racial and social predictors of longitudinal cervical measures: the Cervical Ultrasound Study. Journal of Perinatology. 2017;37(4):335–339.

108. Bligard K, Temming LA, Stout MJ, et al. 85: Performance of cervical length screening in african american women. American Journal of Obstetrics and Gynecology. 2018;218(1, Supplement):S62–S63.

109. Menon R. Spontaneous preterm birth, a clinical dilemma: Etiologic, pathophysiologic and genetic heterogeneities and racial disparity. Acta Obstetricia et Gynecologica Scandinavica. 2008;87(6):590–600.

110. Menon R, Pearce B, Velez DR, et al. Racial disparity in pathophysiologic pathways of preterm birth based on genetic variants. Reproductive Biology and Endocrinology. 2009;7:62.

111. Koullali B, van Zijl MD, Kazemier BM, et al. The association between parity and spontaneous preterm birth: a population based study. BMC Pregnancy Childbirth. 2020;20(1):233.

